# Quantifying representativeness in randomized clinical trials using machine learning fairness metrics

**DOI:** 10.1101/2021.06.23.21259272

**Authors:** Miao Qi, Owen Cahan, Morgan A. Foreman, Daniel M. Gruen, Amar K. Das, Kristin P. Bennett

**Affiliations:** Rensselaer Polytechnic Institute, Department of Computer Science, Troy, New York, USA; Rensselaer Polytechnic Institute, Department of Mathematical Sciences, Troy, New York, USA; IBM Research, Cambridge, Massachusetts, USA

**Keywords:** population representativeness, machine learning, randomized clinical trials, subgroup, health equity

## Abstract

**Objective:** We formulate population representativeness of randomized clinical trials (RCTs) as a machine learning (ML) fairness problem, derive new representation metrics, and deploy them in visualization tools which help users identify subpopulations that are underrepresented in RCT cohorts with respect to national, community-based or health system target populations.

**Materials and Methods:** We represent RCT cohort enrollment as random binary classification fairness problems, and then show how ML fairness metrics based on enrollment fraction can be efficiently calculated using easily computed rates of subpopulations in RCT cohorts and target populations. We propose standardized versions of these metrics and deploy them in an interactive tool to analyze three RCTs with respect to type-2 diabetes and hypertension target populations in the National Health and Nutrition Examination Survey (NHANES).

**Results:** We demonstrate how the proposed metrics and associated statistics enable users to rapidly examine representativeness of all subpopulations in the RCT defined by a set of categorical traits (e.g., sex, race, ethnicity, smoker status, and blood pressure) with respect to target populations.

**Discussion:** The normalized metrics provide an intuitive standardized scale for evaluating representation across subgroups, which may have vastly different enrollment fractions and rates in RCT study cohorts. The metrics are beneficial complements to other approaches (e.g., enrollment fractions and GIST) used to identify generalizability and health equity of RCTs.

**Conclusion:** By quantifying the gaps between RCT and target populations, the proposed methods can support generalizability evaluation of existing RCT cohorts, enrollment target decisions for new RCTs, and monitoring of RCT recruitment, ultimately contributing to more equitable public health outcomes.

## BACKGROUND AND SIGNIFICANCE

Inequitable representation and evaluation of diverse subgroups in randomized clinical trials (RCTs) and other clinical research may generate unfair and avoidable differences in population health outcomes^1-4^. In an analysis of trials conducted by Pfizer between 2011 and 2020, scientists found an urgent need for solutions to enhance diverse representation across all populations within clinical research^5^. Health inequity attracted great public attention during the COVID-19 pandemic^6-8^. For example, race and ethnicity are identified factors associated with risk for COVID-19 infection and mortality^9-11^. The adequate enrollment of participants with diverse race and ethnicity is required in clinical trials to ensure valid treatment effect conclusions and to support reliable generalizability of clinical trial results across subpopulations.

A well-designed RCT is considered the most reliable way to estimate cause-effect relationships between treatments and outcomes^12,13^. The randomization process, which makes RCTs gold standards of treatment effectiveness, contains two random assignments, one from target population to trial cohort and the other from trial cohort to different experimental groups^14,15^. The first random assignment is critical to the applicability and generalizability of clinical findings^16-18^ but has received much less attention than the second one. Figure 1 demonstrates that if a latent patient trait guides the patient assignment into the study and affects the outcome, then the study generalizability to other reference populations may be limited from a causal inference perspective.

**Figure 1.**
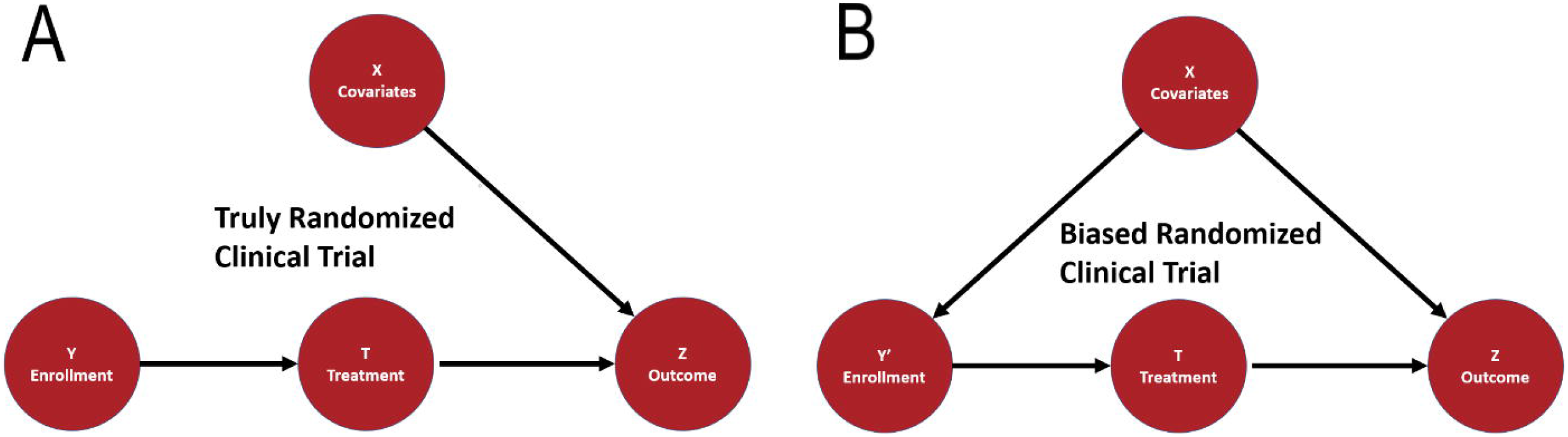
The causal models of truly randomized clinical trials and biased randomized clinical trials. *X* represents the subject covariates; *Y* is the assignment of a subject to a trial; *T* indicates the treatment; and *Z* is the outcome. The black arrows represent causal dependencies between variables. **A**. In the causal model for truly randomized clinical trials, no dependency should exist between *X* and *Y*. Thus, the observed probability of outcome *Z* given the treatment is a good estimate of whether the treatment causes the outcome. **B**. In the causal model for biased randomized clinical trials, an arrow exists between *X* and *Y’*, which indicates the dependence. Thus, invalid causal inferences may be estimated for treatment efficacy among some subpopulations and result in unfair and avoidable population health disparities.

### Population representativeness and previous works

Given the target population (i.e., the broad group of people to which RCT results are intended to apply) of a study on any disease, metrics have been developed to determine if a subpopulation (i.e., a subset of the target population that share a single or multiple common traits and thus can be distinguished from the rest) is underrepresented in the study cohort. Examples of traits include demographics, socioeconomic status, and clinical characteristics.

Existing metrics and measures of population representativeness are generally of two types: (1) comparing rates of subgroups enrolled in a clinical trial versus those in the target population (i.e. enrollment fraction, EF); (2) comparing rates of eligible patients according to inclusion and exclusion criteria versus those in target patient populations (i.e. GIST 2.0).

EF is a widely used measure of participation disparities in clinical trials. For a given disease, it is defined as the number of trial participants divided by the estimated US cases in each subgroup^19,20^. EF is usually a very small number by definition and requires the total number of target population for calculation, which makes potential underrepresentation and discriminations due to subgroup membership with respect to EF challenging to compare and calculate in a numerically stable way. We prove that our proposed metrics based on EF can be obtained through easily calculated and more intuitive rates. Researchers typically assess subgroup representativeness by comparing subgroup EFs with that of a reference subgroup (e.g., non-Hispanic White individuals) that is traditionally advantaged. Our method calculates possible underrepresentation of all subgroups at the same time.

Complex but valuable representativeness metrics like GIST 1.0 and 2.0^21,22^ calculate the generalizability of clinical studies by comparison of eligible populations with target populations based on electronic healthcare records and evaluate the restrictiveness of trial eligibility criteria. Our proposed metrics also compare rates between trial cohorts and target populations, but deal with multiple traits in the cohort differently. GIST calculates measures of each trait and then obtains a final score from the univariate trait measures. We instead calculate representation metrics for all possible subgroups created by the multiple traits and then focus on visualizations and statistical methods that enable users to effectively identify significantly underrepresented subgroups with respect to the target populations. By indicating the representativeness of all possible subgroups, our approach could eventually be combined with GIST-type approaches to help illuminate the “black box” of sample selection and trial generalizability in a single trial and across multiple trials.

### Machine learning fairness and previous works

Machine Learning (ML) Fairness metrics have been developed to quantify and mitigate bias in ML and AI models^23-25^. To improve the performance of existing RCT representativeness measurements, we consider assignment to the RCT a random binary classification problem and develop standardized metrics for RCTs based on variations of ML fairness metrics by mapping to the context of RCTs. ML fairness metrics quantify potential bias towards protected groups in trained ML classification model outcomes. Our metrics, instead of comparing positive and negative classes based on model outcomes, focus on the trial-subject data generation process within the RCT. Our novel insight is to regard subject assignment to an RCT as a classification function that is random and then create variants of ML fairness metrics.

Our metrics capture how well the actual assignment of subjects to an RCT cohort matches with a truly random assignment. The statistical properties of the hypothetical random assignment from a target population can be estimated using community-based or nationally representative datasets of individual characteristics, such as the National Health and Nutrition Examination Survey (NHANES)^26^ or from electronic medical records (EMR).

### Consolidated Standards of Reporting Trials and previous works

Our main goal is to eliminate or reduce inequitable representations in the subject enrollment stage by measuring and identifying equity gaps which persist across different subpopulations. Our method augments the Consolidated Standards of Reporting Trials (CONSORT)^27,28^ statement and its extension CONSORT-Equity^29^, which aims to avoid biased results from incomplete or nontransparent research reports that could mislead decision-making in healthcare. Our method supports incorporating representativeness evaluation before, during, and after any RCTs. Additionally, it can help Institutional Review Board (IRB) better evaluate the equity in trial-design stages and assist FDA regulators to ensure a fair distribution of clinical benefits to both the study sample and the general population.

Our proposed representativeness metrics are expected to identify subgroups that are insufficiently recruited into and represented in the clinical trial cohort using study summary data only, ensuring privacy, security, and confidentiality of health information. These metrics can then be used by clinicians, clinical researchers, and health policy advocates to assess potential gaps in the applicability of clinical trials in real-world settings.

### Our contributions

The contributions discussed in this paper are: (1) Formulating the problem of representativeness evaluation in RCTs as a comparison between a truly random assignment function and the actual assignment observed in the clinical trial cohort; (2) Deriving new metrics for representativeness of RCT based on ML fairness metrics; (3) Utilizing proposed metrics to measure subject representation of RCT cohorts with respect to a target population; (4) Identifying needs, gaps, and barriers of equitable representation of various subgroups in RCTs; (5) Designing a tool (an R-Shiny App) to automatically evaluate trial representativeness through on-demand subject stratification and distribute reports containing visualizations and explanations for different users.

## METHODS AND MATERIALS

We establish a general mapping from RCT to ML Fairness and then derive metrics to evaluate the population representation of RCTs based on ML fairness measures^30-36^. We provide a visual representation of results with associated statistical tests to transparently communicate the quantitative results to diverse user groups.

Table 1 provides a glossary of fairness and representativeness terms used throughout the manuscript.

**Table 1.**
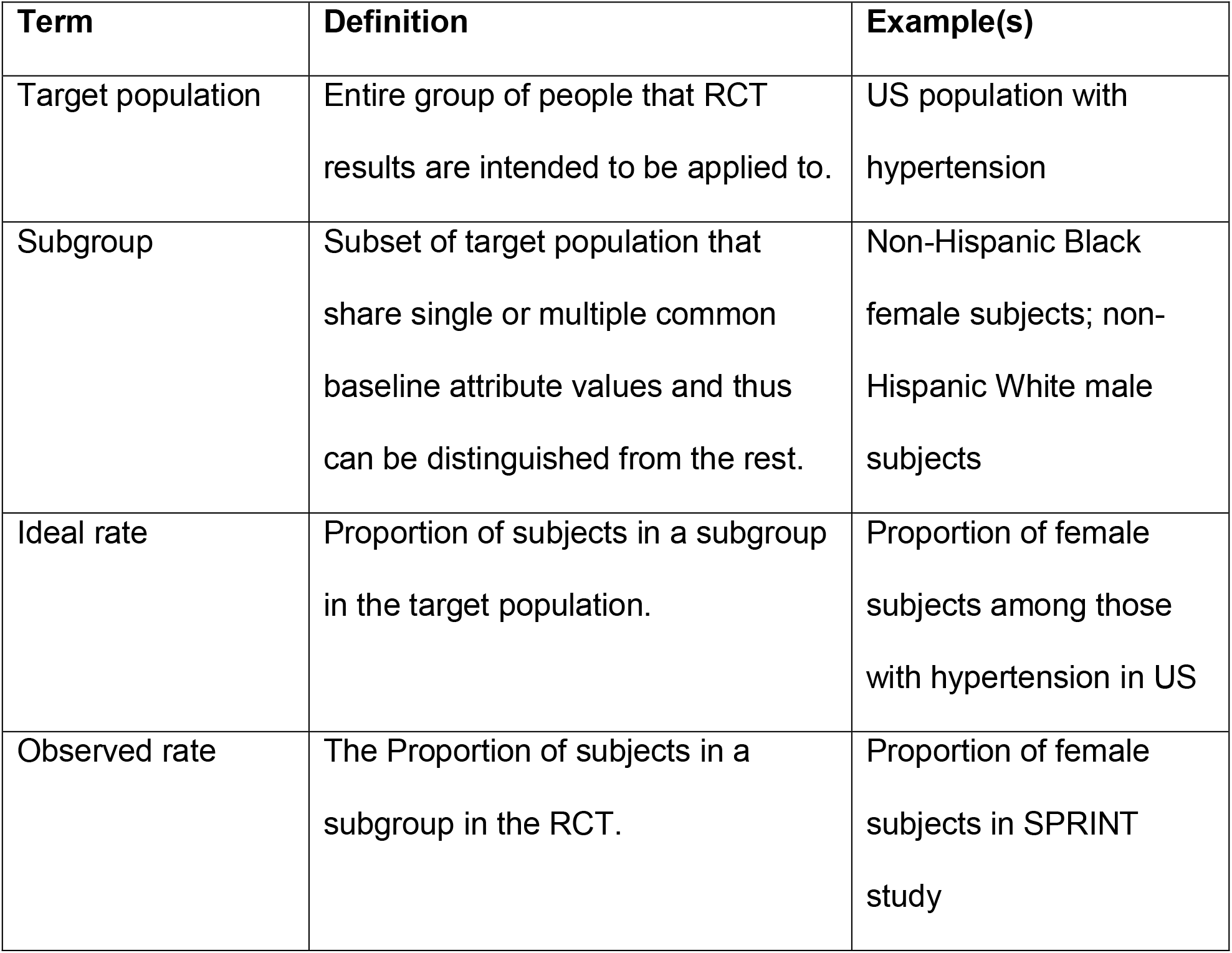
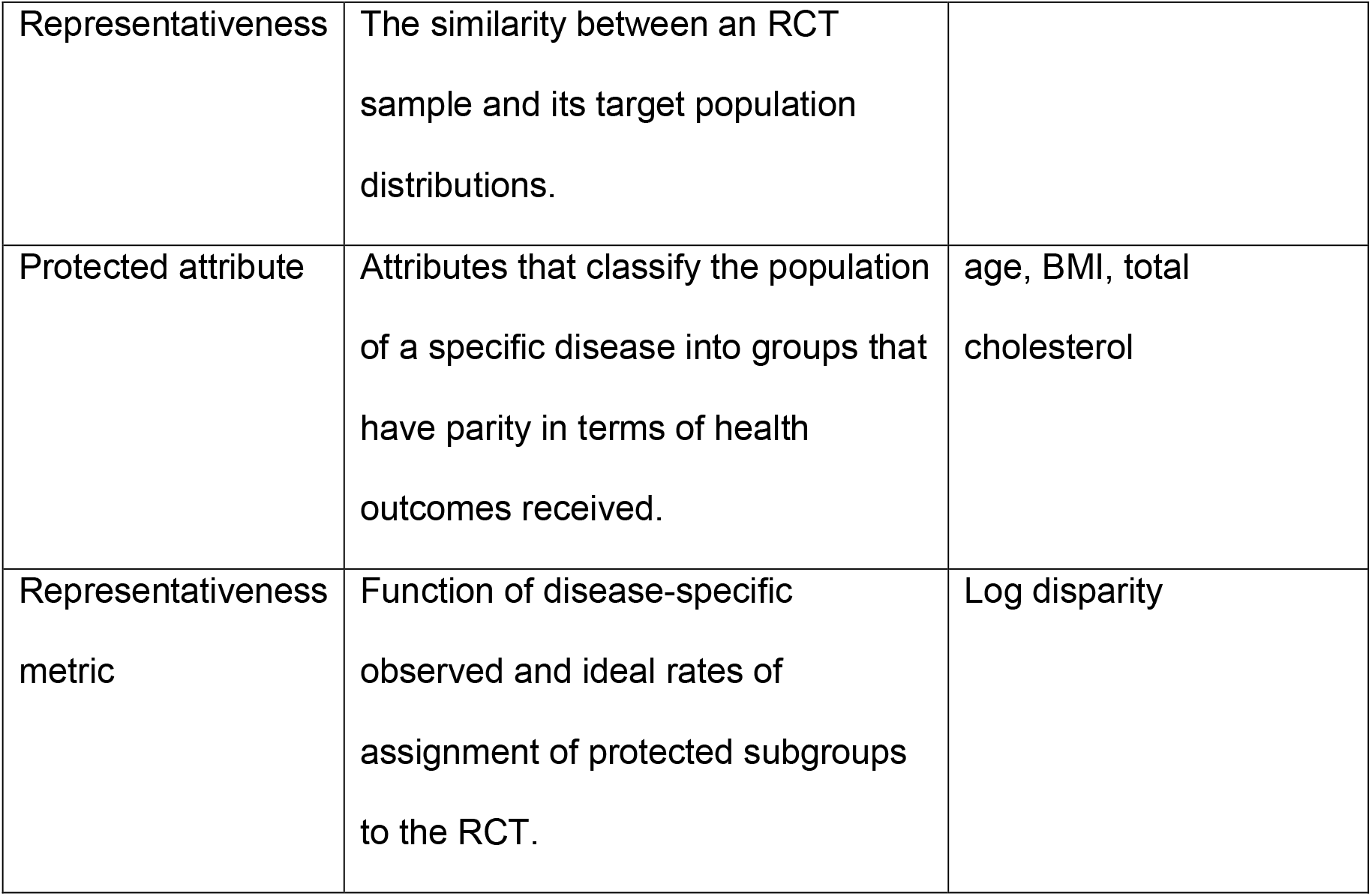
Glossary

| **Term** | **Definition** | **Example(s)** |
| --- | --- | --- |
| Target population | Entire group of people that RCT results are intended to be applied to. | US population with hypertension |
| Subgroup | Subset of target population that share single or multiple common baseline attribute values and thus can be distinguished from the rest. | Non-Hispanic Black female subjects; non-Hispanic White male subjects |
| Ideal rate | Proportion of subjects in a subgroup in the target population. | Proportion of female subjects among those with hypertension in US |
| Observed rate | The Proportion of subjects in a subgroup in the RCT. | Proportion of female subjects in SPRINT study |
| Representativeness | The similarity between an RCT sample and its target population distributions. |  |
| Protected attribute | Attributes that classify the population of a specific disease into groups that have parity in terms of health outcomes received. | age, BMI, total cholesterol |
| Representativeness metric | Function of disease-specific observed and ideal rates of assignment of protected subgroups to the RCT. | Log disparity |

### RCT representativeness and ML fairness

In an ML prediction model, given a feature vector *x* of subject from distribution 𝒫, a binary classifier predicts if the subject is positive (*y*′ = 1) or negative (*y*′ = 0). The true outcome is *y* ∈ {0,1}. Within RCTs, the feature vector *x* is the protected attributes or subject traits; the binary classifier assigns subjects into the study cohort, where *y*′ = 1 means a subject is recruited while *y*′ = 0 means not recruited or exclusion. *y* is the true random assignment result of the subject into the study from the whole target population.

For RCT representativeness evaluation, each available individual (i.e. a person who has the studied disease) is defined by *I* = (*X, y*) = ((*x, x*′), *y*), where *x* ∈ *X* represents the protected attributes, *x*′ ∈ *X* represents the unprotected attributes, and *y* ∈ {0,1} is the ideal assignment of the individual by a randomized clinical trial. An ideal RCT enrolls subjects i.i.d. from the target population 𝒫. The RCT recruitment strategy can be treated as a binary classifier 𝒟(*X*) = *y*′ ∈ {0,1}, denoting the real observed decision induced by 𝒟 on an individual *i*. The subgroups are defined via a family of indicator functions 𝒢. For each *g* ∈ 𝒢, *g*(*x*) = 1 means that an individual with protected attributes *x* is in the subgroup. We utilize protected attributes of three types: demographic characteristics, risk factors and laboratory results.

ML fairness metrics are concerned with guaranteeing similarity results across different subgroups^37^. We assume that the ideal RCT achieves statistical parity^38^, i.e. subgroups are independent of outcomes (*g*(*x*) ⊥ *y*). Then we create metrics based on ML fairness measures of statistical parity violations. The proposed metrics also assume that the ideal assignment of a subject to the RCT and the observed assignment are independent (*y* ⊥ *y*′), and the sizes and the rates of an ideal RCT and the observed trial are the same (*P*(*y* = 1) = *P*(*y*′ = 1)).

The ideal and observed rates of a subgroup are *P*(*g*(*x*) = *s*|*y* = 1) and *P*(*g*(*x*) = *s*|*y*′ = 1), respectively. The enrollment fraction of a subgroup is *P*(*y*′ = 1|*g*(*x*) = *s*). We note by independence assumptions of ideal RCT, *P*(*y*′ = 1|*y* = 1, *g*(*x*) = *s*) = *P*(*y*′ = 1|*g*(*x*) = *s*).

### Log Disparity Metric for RCT

In ML fairness, the disparate impact measure is the ratio of positive rates of both unprotected and protected groups^39^:

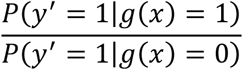

Disparate impact adopts the “80 percent rule” suggested by the US Equal Employment Opportunity Commission (EEOC)^40^ to decide when the result is unfair:

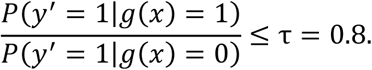

The “80 percent rule” requires the selection rate of a subgroup to be at least 80% of the selection rate of the other subgroups.

As shown in the following theorem, when applied to the RCT, disparate impact reduces to an intuitive quantity based on the enrollment odds of a protected group and in the target.

#### Theorem 1

RCT version of Disparate Impact Metric

Based on the ideal RCT assumptions above, the disparate Impact metric is equivalent to the ratio of enrollment odds of subjects of the protected group in the observed cohort to the ratio of the odds of subjects in the ideal cohort:

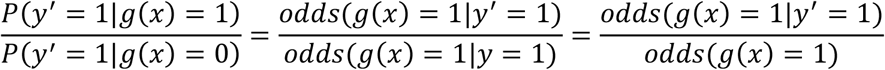

See supplementary materials for proof.

Since log odds provides advantages for ease of understanding, we propose the following metric for RCT.

#### Proposed Metric 1.

The *Log Disparity* metric for measuring how representative of subgroup *g*(*x*) = 1 in observed trial *y′* as compared to ideal population *y* is

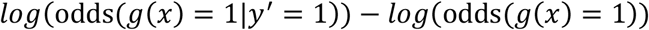

In the log disparity metric, a value of 0 indicates perfect clinical equity. A value smaller than the lower threshold, −τ_*lower*_, implies a potential underrepresentation of a subgroup while a value greater than implies a potential overrepresentation. We further add an upper threshold, τ_*upper*_. A value less than −τ_*upper*_ implies highly underrepresentation; similarly, a value greater than τ_*upper*_ implies highly overrepresentation. Values between −τ_*lower*_ and τ_*lower*_ mean equitable representation.

Our metric thresholds are selected based on guidance from literature^23,41-43^, but other optimal thresholds under different criteria are allowed as inputs. We use a significance level of 0.05, a lower threshold of -log (0.8), and an upper threshold of -log (0.6).

### Normalized Parity Metric

The ML fairness Equal Opportunity^44^ metric which requires subgroups to have the same true positive rates can also be applied to RCTs.

#### Theorem 2

RCT version of Equal Opportunity Metric

Let ideal RCT assumptions hold and *g*(*x*) be binomial random variable, then the ML fairness Equal Opportunity metric has the following equivalent form:

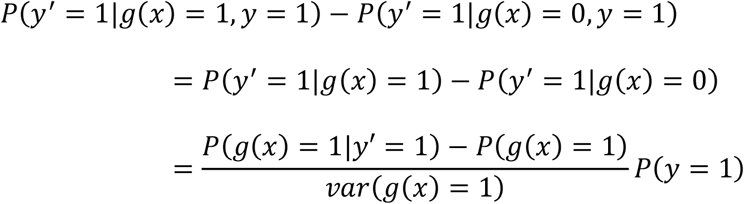

See supplementary materials for proof. The proportion of population in the trial, *P*(*y* = 1), is extremely small and not very meaningful, thus we propose a new metric. The *Normalized Parity* metric measures the difference in rates of protected group in the trial and in the population scaled by the variance of the protected group in the target population.

#### Proposed Metric 2.

The *Normalized Parity* metric for measuring how representative of subgroup *g*(*x*) = 1 in observed trial *y*′ as compared to ideal population *y*

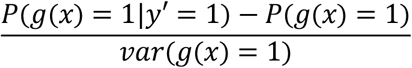

The proposed Log Disparity and Normalized Parity metrics have several nice properties.

1. They are easy to compute. The observed rates of each subgroup, *P*(*g*(*x*) = 1|*y*′ = 1), are estimated from trial data. The ideal rates and variance, *P*(*g*(*x*)) and *var*(*g*(*x*)), are estimated for the desired target population 𝒫 using surveillance datasets such as NHANES or EMR. The required estimates are robust to missing data. Individual privacy can be protected since only summary statistics are required for the proposed metrics, avoiding the pitfalls of alternative metrics requiring per subject calculations^45^.
2. Both metrics have a common interpretation for subgroups with very different background rates: 0 means that demographic parity holds, <0 means subgroup is underrepresented, and >0 means subgroup is overrepresented.
3. Statistical tests quantify the significance of observed disparities for each subgroup which take into account the RCT study size and estimation errors of the ideal assignment rate. We use a one-proportion two-tailed z-test to determine whether the observed rate is significantly deviated from the ideal population rate. We use Benjamini-Hochberg to correct for multiple comparisons across all subgroups. If the difference between observed and ideal rates is not statistically significant, the subgroup is treated as representative; otherwise, we will use metrics to quantify the subgroup representativeness. Other statistical tests could be used. See supplement for details.

### RCT trial data

We assess the proposed methodologies on three real-world RCTs: ACCORD^46^, ALLHAT^47^, and SPRINT^48^ in BioLINCC with the ideal subgroup assignment rate calculated from individuals with matched disease conditions in NHANES. According to participants’ baseline characteristics typically summarized in Table 1s of clinical trial reports, we selected nine protected attributes. We categorize continuous variables based on the CDC-approved standards. Subject data obtained from RCTs are mapped to the existing NHANES categories. The protected attributes examined here are (a) demographic characteristics (gender, race/ethnicity, age, and education); (b) baseline risk factors (smoking status, body mass index (BMI), and systolic blood pressure (SBP)); (c) baseline laboratory test results (fasting glucose (FG) and total cholesterol (TC)).

The observed rates of the subgroup are calculated from the RCT data

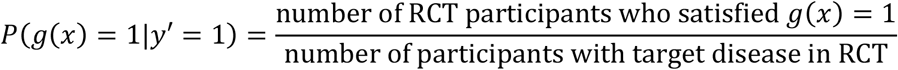

For each study, we construct all possible subgroups that can be instantiated as *g*(*x*). We define 29 univariate, 109 bivariate, and 306 multivariate subgroups based on nine protected attributes. In general, any baseline subject attributes can be selected as protected attributes in our approach.

### Target population

In our experiment, ideal rates from target populations (*P*(*g*(*x*) = 1|*y* = 1)) are calculated from NHANES 2015-2016 using the R survey() package^49^ which accounts for potential bias from complex survey designs. The NHANES population selected varies based on study objectives and desired target population. To evaluate ACCORD^46^, we estimate ideal rates of subgroups of diabetic individuals in the US using subjects who report having diabetes in NHANES, and we use subjects who report having hypertension in NHANES as the target population to evaluate ALLHAT^47^ and SPRINT^48^. These criteria could be modified to consider study inclusion and exclusion criteria depending on the goals of analysis.

Since users may have better target population data that match their studies, user-provided target population datasets and multiple target files are allowed. For example, clinicians who focus on their local communities could use the community or health-system population as the target to evaluate the equity of RCTs, whereas researchers who work on a global disease, the target population may be better estimated from global population datasets.

## RESULTS

To demonstrate the proposed metric, we created a visualization using different colors to represent different representativeness levels in RCTs. For compact presentation, we focus on the log disparity metric. Figure 2 illustrates how the log disparity function applies to relative common subgroups Female and Female Non-Hispanic Black in ACCORD.

**Figure 2.**
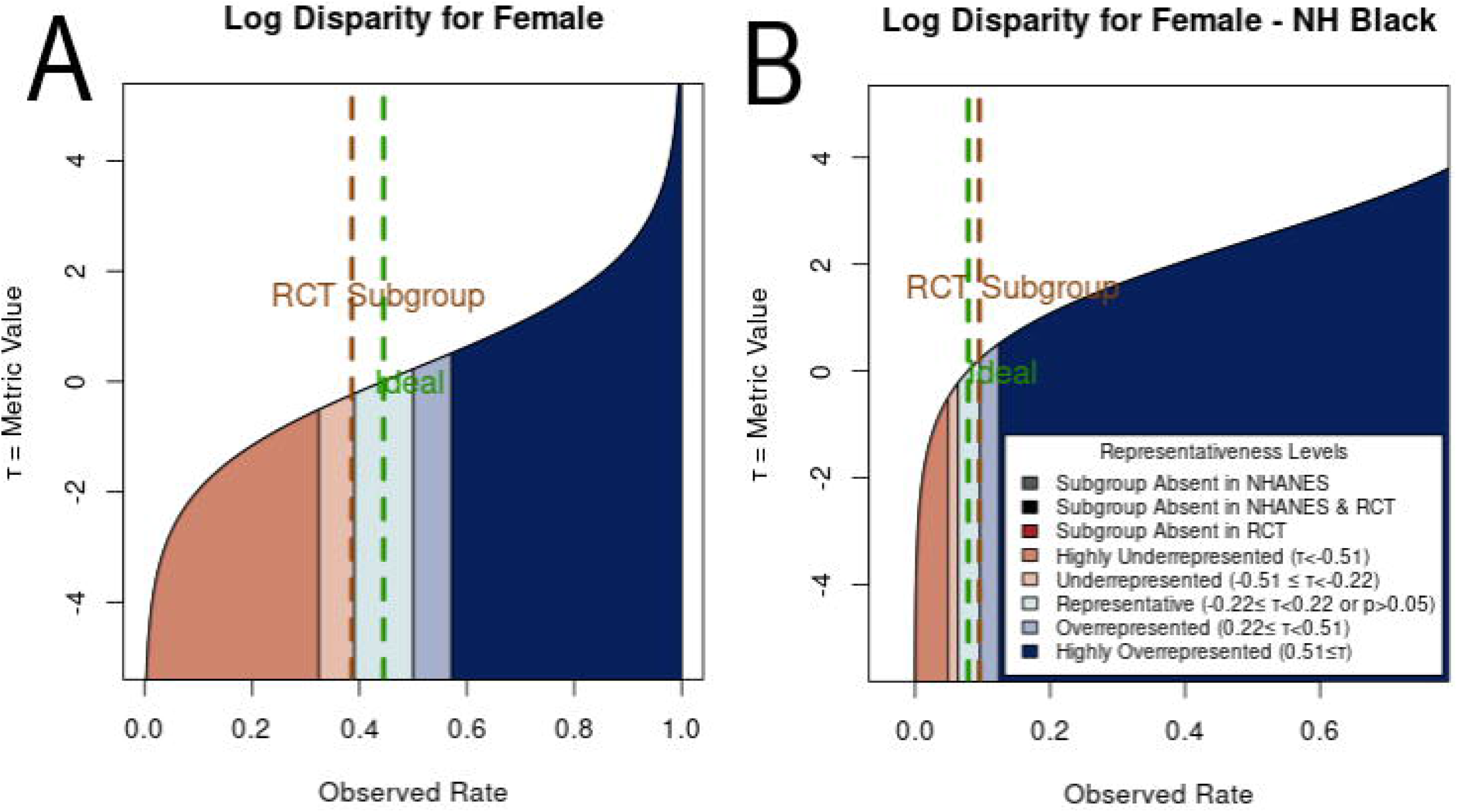
The shift of representativeness distribution of Log Disparity metric for different patient subgroups with type-2 diabetes in ACCORD. The green line corresponds to the ideal rate for the subgroup determined from NHANES. The brown line indicates the rate actually observed. **A**. Log Disparity as function of observed rate for female subgroup. **B**. Log Disparity as function of observed rate for female non-Hispanic black subgroup.

As shown in Figure 2A, for women with type-2 diabetes, the ideal rate from NHANES is 0.445 while the observed RCT rate is 0.386. The observed female-subject rate falls into the light orange region, which reveals the underrepresentation of female subjects. For Figure 2B, when the subgroup of interest is changed to non-Hispanic black female participants, the ideal rate decreases to 0.079 and the observed rate becomes 0.095. Now the interested subgroup falls into the teal region, which means that non-Hispanic black female participants are equitably represented in ACCORD. This indicates the influence of protected attribute race/ethnicity on the representativeness evaluation. By comparing Figure 2A and 2B, we can observe that metric functions change as the ideal rate changes.

The representativeness of 29 univariate subgroups for three RCTs are shown in Table 2 and 3. Dark red represents the subgroups absent from the RCT; light orange and orange red indicate that subgroups are underrepresented or highly underrepresented in the RCT relative to the target population; light and dark blue specify the potentially overrepresented or highly overrepresented subgroups; teal shows the subgroup is either equitably represented or has no significant difference; grey indicates that no individuals with selected protected attributes exist in estimated target population; black indicates absent subgroup in both estimated target population and RCT.

**Table 2.** Representativeness of subgroups defined by a single protected attribute using Log Disparity for three real-world RCTs. Subgroups are defined by demographic characteristics. Black cells labeled with “NA” indicate that a specific category/value was not measured in the RCT; teal cells with a star indicates that no statistically significant difference between subgroups from the RCT and target population. Ages are in years.

| Demographic Characteristics | Representativeness |  |  |
| --- | --- | --- | --- |
|  | ACCORD | ALLHAT | SPRINT |
| **Gender** |  |  |  |
| Female | -0.25 | -0.21 | -0.68 |
| Male | 0.25 | 0.21 | 0.68 |
| **Age (Diabetes/Hypertension)** |  |  |  |
| 18-44/18-39 | -4.21 | -Inf | -Inf |
| 45-64/40-59 | 0.86 | -0.87 | -0.74 |
| 64+/59+ | -0.28 | 1.45 | 1.32 |
| **Race/Ethnicity** |  |  |  |
| Non-Hispanic White | 0.23 | -0.73 | -0.30 |
| Non-Hispanic Black | 0.32 | 1.04 | 0.96 |
| Non-Hispanic Asian | NA | -1.36 | -1.63 |
| Hispanic | -1.07 | 0.53 | -0.16 |
| Other/Unknown | 0.14 | -1.58 | -1.90 |
| **Education** |  |  |  |
| Less than high school | -0.46 | 0.95 | -2.32 |
| High-school graduate | 0.30 | 0.14 | -0.48 |
| Some college/Technical school | -0.06 | -0.38 | 0.31 |
| College degree or higher | 0.14 | -1.83 | 0.69 |

**Table 3.** Representativeness of subgroups defined by a single protected attribute using Log Disparity for three real-world RCTs. Subgroups are defined by clinical characteristics. Systolic blood pressure unit = mm Hg; Fasting glucose unit = mmol/L.

| Clinical Characteristics | Representativeness |  |  |
| --- | --- | --- | --- |
|  | ACCORD | ALLHAT | SPRINT |
| **Cigarette-smoking status** |  |  |  |
| Current smoker | -0.91 | -0.93 | -1.49 |
| Not smoke | 0.91 | 0.93 | 1.47 |
| **Body-mass index group** |  |  |  |
| Underweight | -2.57 | 0.04★ | -1.03 |
| Normal weight | -0.26 | 0.21 | -0.56 |
| Overweight | 0.06 | 0.37 | -0.17 |
| Obese | 0.05 | -0.51 | -0.79 |
| **Systolic blood pressure** |  |  |  |
| <120 | -0.94 | -1.95 | -Inf |
| 120-129 | -0.29 | -1.13 | -Inf |
| 130-139 | 0.52 | -0.19 | 0.87 |
| >=140 | 0.52 | 1.57 | 1.29 |
| **Total cholesterol** |  |  |  |
| Normal | -0.26 | -1.07 | 0.04★ |
| High | 0.24 | 0.87 | -0.19 |
| **Fasting glucose** |  |  |  |
| <5.6 | -0.56 | 0.42 | 1.17 |
| 5.6-6.9 | -0.85 | -1.71 | -0.66 |
| >=7 | 0.81 | 0.06 | -2.17 |

We evaluate our ideal estimates for ACCORD, ALLHAT, and SPRINT using prior literature. For example, an estimated probability of female patients among US hypertensive population in 2015^50^, calculated through Bayes’ formula, is about 47%. Comparing to the summary statistics in published literature (i.e., about 47% subjects are women in ALLHAT and 36% subjects are women in SPRINT^51-54^), ALLHAT captures the gender distribution among real-world hypertensive participants while SPRINT fails to enroll enough female participants.

The color change across categories of an attribute highlights interesting trends in subject representation. Among three studies, only two attributes achieved equitable representation across all subgroups: gender in ALLHAT and TC in SPRINT. From the tables, we observe that current smokers, young participants, non-Hispanic Asian subjects, subjects with SBP under 130 mm Hg or FG between 5.6-6.9 mmol/L are frequently underrepresented. This indicates that some subgroups in the target population are missing or inadequately represented in the RCTs. The decision-making on a subject, e.g. aged 40, based on the SPRINT study would require additional evidence beyond this study. Also, participants with lower education levels tend to be more underrepresented in the SPRINT while participants with higher education levels tend to be more underrepresented in the ALLHAT. This points out that potential social determinant confounders may exist in the RCT. We note, across all three studies, non-Hispanic black participants are overrepresented, perhaps reflecting efforts to ensure minority participation or reflecting study locations. In both hypertension RCTs, Asian subjects may have been insufficiently enrolled. This underrepresentation may also reflect study choices or locations. These trends have to be validated by analysis on more RCTs.

For subgroups defined by multiple attributes, sunburst plots better visualize the change of subgroup representation by adding additional protected attributes, as shown in Figure 3. For each type of protected attributes (i.e., demographic characteristics, risk factors, and lab results), separate sunburst charts are generated since their matched population from NHANES are different.

**Figure 3.**
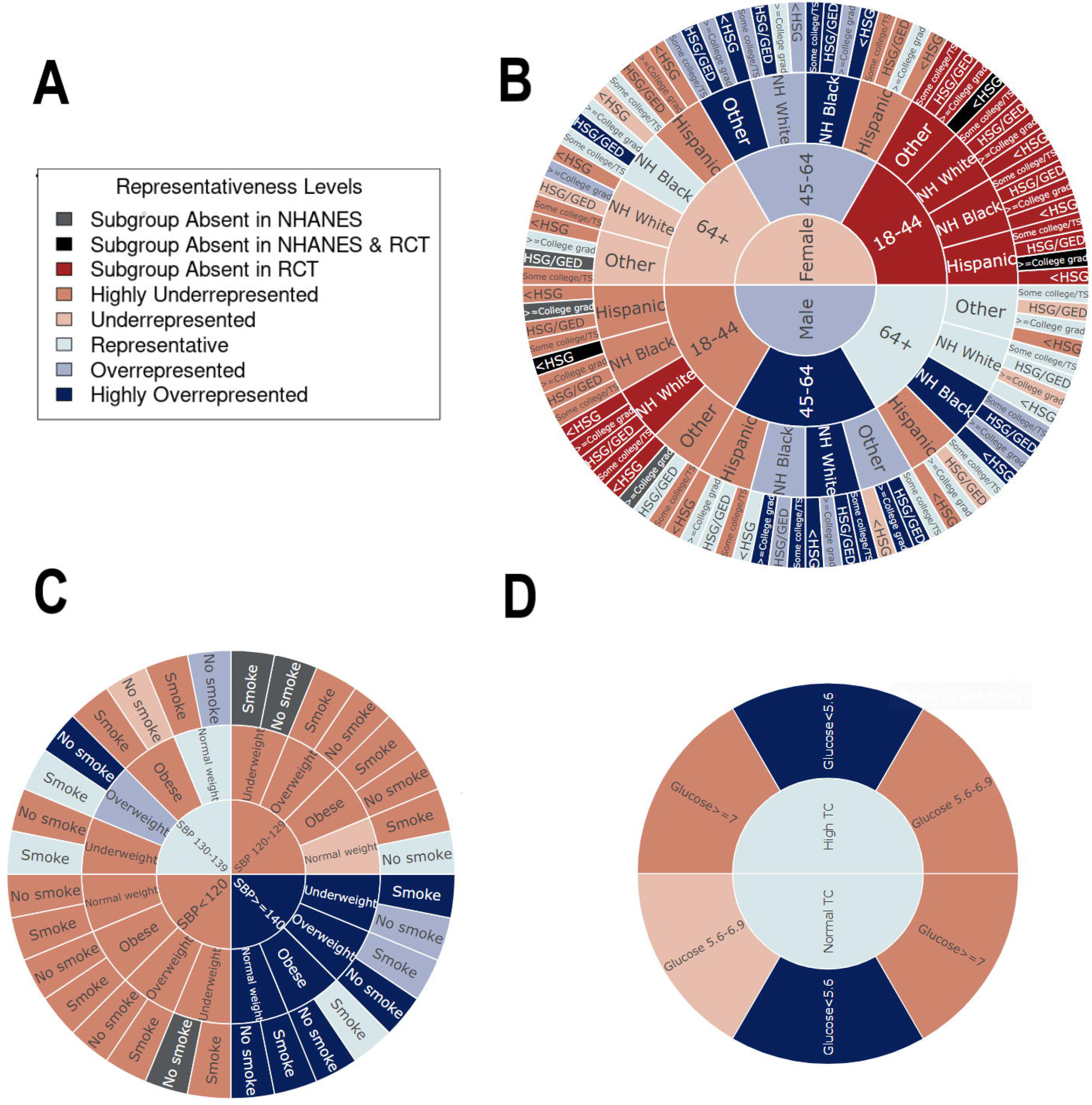
Representativeness results measured by Log Disparity. **A**. Color code of representativeness levels. **B**. Representativeness of ACCORD RCT subgroups in sunburst plot with inner to outer rings defined by demographic characteristics gender, age, race/ethnicity, and education level respectively. **C**. Representativeness of ALLHAT RCT subgroups in sunburst plot with inner to outer rings defined by risk factors SBP, BMI, and smoking status respectively. **D**. Representativeness of SPRINT RCT subgroups in sunburst plot with inner to outer rings defined by lab results total cholesterol and fasting glucose respectively.

Figure 3 demonstrates log disparity results for ACCORD on demographic characteristics, ALLHAT on risk factors, and SPRINT on lab results. The interactive sunburst diagram enables users to investigate many subgroups simultaneously to identify missing or underrepresented subgroups in RCTs and NHANES. For example, young female subjects aged under 45 are missing entirely. As shown in Figure 3D, with an additional attribute FG, new subgroups such as participants with glucose>=7 mmol/L are highly underrepresented for both high and normal TC. This indicates the importance of multivariable subgroup analyses in representativeness. Note that underrepresentativeness may be due to legitimate choices in the study inclusion and exclusion criteria. If desired by the user, absent subgroups in NHANES or any target populations can be estimated using smoothing techniques.

The sunburst plots explicitly address diversity, equity, and inclusion of clinical studies with respect to the target population. For instance, Figure 3B identifies the missing evidence in subgroups including any female and non-Hispanic male subjects aged under 45. This lack of subject diversity may lead to similar results as shown for the effectiveness of Actemra on COVID-19 patients, in which the study results flipped after including more marginalized participants. Furthermore, our visualization automatically checks if the inclusion and exclusion criteria are met. Based on the criteria of SPRINT, it successfully excluded subjects with SBP under 130 mm Hg but subjects with potential impaired glucose or diabetes still existed based on the lab results.

## DISCUSSION

An advantage of the proposed metrics is they provide a standardized scale for judging trial representativeness for subgroups with vastly different expected rates in the trial; for example, the estimated ideal rate of participation in the type-2 diabetes trial estimated from NHANES for subgroups of female subjects, female subjects aged over 64, Hispanic female subjects aged over 64, and Hispanic female subjects aged over 64 with high school degree are 0.445, 0.172, 0.025, and 0.006 respectively. Evaluating differences between simple rates for many subpopulations would be more challenging.

To facilitate visualizations of measured performance on clinical trials, we have incorporated a comprehensive set of fairness metrics into our prototype representativeness visualization tool using R shiny to enable researchers and clinicians to rapidly visualize and assess all potential misrepresentation in a given RCT for all possible subgroups. In our application, the number and order of the attributes for the sunburst can be changed by users; for example, instead of Figure 3B, users can visualize representativeness of subgroups for Age with further divisions by Gender and then Race/Ethnicity. With these metrics, users can rapidly determine underrepresentation of subgroups which can serve as basis for determining any limitations of the RCT. The metrics and visualizations can potentially help support evaluation of representativeness of existing RCTs, design of new RCTs, and monitoring of recruitment in ongoing RCTs. The visualization may also help healthcare providers quickly understand the applicability of RCT results to a patient in a subgroup.

Clinical trials are a key component of health equity. In the context of trial equity, underrepresentation or exclusions of disadvantaged participants may reduce their opportunities to live healthy lives. These metrics can also be applied to many types of clinical research and representativeness problems by appropriately adjusting the target population statistics based on the population of interest. Besides use with RCTs, these metrics can be easily modified to assess and visualize any disparities related to health including the distribution of medical care and different levels of living and working conditions for patients if the matching background information is available to obtain the ideal rate of each subgroup. Furthermore, our approach can be used as a frame of reference to guide the clinicians and policy-makers to make decisions with legitimate reasons and evidence. We offer user selections to dynamically control different conditions including subgroup characteristics, metric types, metric cutoffs, under which the users will make their own decisions.

The technical challenges we encountered include determining how to appropriately treat continuous variables such as age and consider inclusion and exclusion criteria when mapping RCT cohorts and NHANES sample population. Currently, we discretize all continuous variables, with alternative approaches left as future work. It may be desirable to further refine the target populations to adjust for missing and underrepresented subgroups due to RCT inclusion and exclusion criteria. We plan to validate our metrics by applying them to more trials and compare results with other metrics such as GIST 2.0. It can also be useful to create a method combining the proposed metrics with GIST to enable detailed subpopulation analyses of inclusion and exclusion criteria and analysis of multiple trials.

## CONCLUSION

Quantifying representation is important for scientific rigor and to build true equity into research designs and methods. Health equity is not just a clinical issue; it is also a socioeconomic concern with broad consequences^55-57^. We developed metrics and methods to evaluate how equitably subgroups are represented in RCTs. Unlike most existing studies which focus on one protected attribute each time (e.g. race) for a single disease (e.g. type-2 diabetes), our proposed approach can analyze clinical trials designed for several diseases such as hypertension and type-2 diabetes, simultaneously and can additionally report representativeness of subgroups defined by multiple attributes including age and race/ethnicity. Our next steps are to utilize these metrics to monitor existing RCTs, help design new RCTs, and provide tools for disseminating findings to different user groups, such as patients, clinicians, data scientists, and policy-makers, who will bring the discoveries into play to advance health equity.

## Data Availability

The example ideal national patient data are calculated from the National Health and Nutrition Examination Survey (NHANES) 2015-2016 conducted by the National Center for Health Statistics (NCHS). The clinical trial data that support the findings of this study are available from Biologic Specimen and Data Repository Information Coordinating Center (BioLINCC) but restrictions apply to the availability of these data, which were used under license for the current study, and so are not publicly available. Data are however available with permission of BioLINCC. The data generated during and analyzed during the current study are available in the GitHub repository, https://github.com/TheRensselaerIDEA/ClinicalTrialEquity.

## Abbreviations

(RCT): Randomized Clinical Trial
(ML): Machine Leaning
(NHANES): National Health and Nutrition Examination Survey
(ACCORD): Action to Control Cardiovascular Risk in Diabetes
(ALLHAT): Antihypertensive and Lipid-Lowering Treatment to Prevent Heart Attack Trial
(SPRINT): Systolic Blood Pressure Intervention Trial

## ETHICAL APPROVAL

This manuscript was prepared using ACCORD, ALLHAT, and SPRINT Research Materials obtained from the NHLBI Biologic Specimen and Data Repository Information Coordinating Center and does not necessarily reflect the opinions or views of the ACCORD, ALLHAT, SPRINT or the NHLBI. All methods were carried out following the NHLBI approved research plan: Equity in Clinical Trials, and all procedures were carried out in accordance with the applicable guidelines and regulations from NHLBI Research Materials Distribution Agreement. The procedures were approved by The Rensselaer IRB as IRB Review Not Required. Informed consent was obtained from all subjects by NHLBI. Data from research participants who refused to permit the sharing of their data are deleted from the repository data set.

## SUPPLEMENTARY MATERIAL

Supplementary material is available in another document.

## AUTHOR CONTRIBUTIONS

K.P.B., A.K.D., D.M.G., and M.A.F. designed and directed the project. M.Q., K.P.B. and O.C. designed the model and the computational framework. M.Q. performed the experiments, analyzed the results, built the application, and wrote the manuscript in consultation with K.P.B., A.K.D., D.M.G., and M.A.F. All authors reviewed the manuscript.

