## Supplementary Materials for "Quantifying representativeness in randomized clinical trials using machine learning fairness metrics"

We provide more details about the metrics derivations and experimental results.

### SUPPLEMENT 1: METRICS PROOFS AND DERIVIATIONS

For a randomized clinical trial (RCT), an available individual is defined by $I = (X, y) = \big((x, x'), y\big)$, where $x \in \chi$ represents the protected attributes, $x' \in \chi$ represents the unprotected attributes, and $y \in \{0,1\}$ is the ideal assignment of the individual by a RCT. The recruitment strategy is treated as a binary classifier $\mathcal{D}(\chi) = y' \in \{0,1\}$, denoting the real observed decision induced by $\mathcal{D}$ on an individual $I$. The subgroups are defined via a family of indicator functions $\mathcal{G}$. For each $g \in \mathcal{G}$, $g(x)$ = 1 means that an individual with protected attributes $x$ is in the subgroup where all individuals have the same values for $x$.

These derivations are based on the following assumptions:

*Assumption 1:* $y \perp y'$: the ideal assignment of a subject to the RCT and the observed assignment are independent.

*Assumption 2:* $y \perp g(x),\ P(y=1|g(x)=1) = P(y=1|g(x)=0) = P(y=1)$ : the protected attributes and assignment to the ideal RCT are independent of subgroup.

*Assumption 3:* $P(y=1) = P(y'=1)$: the size and the rate of the ideal RCT and the observed trial are the same.

**Log Disparity**

Disparity Impact fairness metric measures the bias led by a selection process which generates significantly different outcomes for different groups. Specifically, it requires the positive rate of both unprotected and protected groups to be similar:

$$\frac{P(y' = 1|g(x) = 1)}{P(y' = 1 \mid g(x) = 0)}\ .$$

Log Disparity is derived from ML Disparity Impact to ensure that the subgroup assignment rates in RCT is similar to those in the matched target population.

Proof:

$$\frac{P(y' = 1|g(x) = 1)}{P(y' = 1|g(x) = 0)} = \frac{\big(P(y' = 1, g(x) = 1)\big)/\big(P(g(x) = 1)\big)}{\big(P(y' = 1, g(x) = 0)\big)/\big(P(g(x) = 0)\big)}$$

$$= \frac{\big(P(g(x) = 1|y' = 1)P(y' = 1)\big)\big(P(g(x) = 0)\big)}{\big(P(g(x) = 0|y' = 1)P(y' = 1)\big)\big(P(g(x) = 1)\big)}$$

$$= \frac{P(g(x) = 1|y' = 1)P(g(x) = 0)}{P(g(x) = 0|y' = 1)P(g(x) = 1)}$$

$$= \frac{P(g(x) = 1|y' = 1)(1 - P(g(x) = 1)|y = 1)}{\big(1 - P(g(x) = 1|y' = 1)\big)P(g(x) = 1|y = 1)} = \frac{odds(g(x) = 1|y' = 1)}{odds(g(x) = 1|y = 1)}$$

$$= \frac{odds(g(x) = 1|y' = 1)}{odds(g(x) = 1)} \blacksquare$$

The above proof produces a ratio of the odds ratios of $P(g(x) = 1|y' = 1)$ and $P(g(x) = 1)$. We simplify the formula by taking the natural logarithm of the ratio, which leads to the difference in log odds

$$log\left(\frac{P(g(x) = 1|y' = 1)}{1 - P(g(x) = 1|y' = 1)}\right) - log\left(\frac{P(g(x) = 1)}{1 - P(g(x) = 1)}\right) \ .$$

For Log Disparity, a value of 0 indicates the perfect clinical representativeness. A value smaller than 0 implies a potential underrepresentation of the participant group while a value greater than 0 implies a potential overrepresentation of the participant group.

Disparity Impact adopts the "80 percent rule" suggested by the US Equal Employment Opportunity Commission (EEOC) to decide when the result is unfair. That is,

$$\frac{P(y' = 1|g(x) = 1)}{P(y' = 1|g(x) = 0)} \leq \tau = 0.8 \ .$$

For Log Disparity, since we take the natural logarithm of the original metric, the new threshold should become $\tau_{RCT} = -log(1 - \tau_{ML})$, where $\tau_{ML} \in [0,1]$ . So, the recommended representativeness threshold of Log Disparity is -log (0.8).

**Normalized Parity Metric**

ML Equal Opportunity requires subgroups to have similar true positive rates: $P(y' = 1|g(x) = 1, y = 1) - P(y' = 1|g(x) = 0, y = 1)$. It emphasizes on the correctly prediction of positive labels but ignores the possible influence of false positive. Normalized Parity is derived from the ML metric Equal Opportunity and ensures that the proportions of people assigned in an ideal RCT but with different protected attribute values should be assigned into the real clinical trial with equal rates. Since the value of $P(y' = 1)$ is extremely small in RCTs, the usual ML metric Equal Opportunity would be very small. Thus, we scale the metric by dividing it by $P(y' = 1)$.

Proof:

$$\frac{P(y' = 1|g(x) = 1, y = 1) - P(y' = 1|g(x) = 0, y = 1)}{P(y' = 1)}$$

$$= \left(\frac{P(y' = 1, g(x) = 1|y = 1)}{P(g(x) = 1|y = 1)} - \frac{P(y' = 1, g(x) = 0|y = 1)}{P(g(x) = 0|y = 1)}\right)\frac{1}{P(y' = 1)}$$

$$= \left(\frac{P(y' = 1, g(x) = 1)}{P(g(x) = 1)} - \frac{P(y' = 1, g(x) = 0)}{P(g(x) = 0)}\right)\frac{1}{P(y' = 1)}$$

$$= \left(\frac{P(g(x) = 1|y' = 1)P(y' = 1)}{P(g(x) = 1)}\right.$$

$$\left. - \frac{P(g(x) = 0|y' = 1)P(y' = 1)}{P(g(x) = 0)}\right)\frac{1}{P(y' = 1)}$$

$$= \left(\frac{P(g(x) = 1|y' = 1)}{P(g(x) = 1)} - \frac{P(g(x) = 0|y' = 1)}{P(g(x) = 0)}\right)\big(P(y' = 1)\big)\frac{1}{P(y' = 1)}$$

$$= \frac{P(g(x) = 1|y' = 1)}{P(g(x) = 1)} - \frac{P(g(x) = 0|y' = 1)}{P(g(x) = 0)}$$

$$= \frac{P(g(x) = 1|y' = 1)}{P(g(x) = 1)} - \frac{1 - P(g(x) = 1|y' = 1)}{1 - P(g(x) = 1)}$$

$$= \frac{1 - P(g(x) = 1)}{1 - P(g(x) = 1)}\,\frac{P(g(x) = 1|y' = 1)}{P(g(x) = 1)}$$

$$- \frac{P(g(x) = 1)}{P(g(x) = 1)}\,\frac{1 - P(g(x) = 1|y' = 1)}{1 - P(g(x) = 1)}$$

$$= \frac{P(g(x) = 1|y' = 1) - P(g(x) = 1)P(g(x) = 1|y' = 1)}{P(g(x) = 1)(1 - P(g(x) = 1))}$$

$$- \frac{P(g(x) = 1) - P(g(x) = 1)P(g(x) = 1|y' = 1)}{P(g(x) = 1)(1 - P(g(x) = 1))}$$

$$= \frac{P(g(x) = 1|y' = 1) - P(g(x) = 1)}{P(g(x) = 1)(1 - P(g(x) = 1))}$$

$$= \frac{P(g(x) = 1|y' = 1) - P(g(x) = 1)}{var(g(x) = 1)} \blacksquare$$

The last transformation comes from the fact that $g(x)$ is a Bernoulli random variable.

Therefore, we get the Normalized Parity metric as

$$\frac{P(g(x) = 1|y' = 1) - P(g(x) = 1)}{var(g(x) = 1)}.$$

The Normalized Parity metric measures the difference in rate of protected group in the trial and the rate of the protected group in the population scaled by the variance of the protected group in the population.

For Normalized Parity, a value of 0 indicates the perfect clinical representativeness. A value smaller than 0 implies a potential underrepresentation of the participant group while a value greater than 0 implies a potential overrepresentation of the participant group. The recommended representativeness threshold of this metric is 0.1, which is suggested by the AI Fairness 360 Open Source Toolkit.

### SUPPLEMENT 2: STATISTICAL ANALYSIS

We want to test if the observed rate of a subgroup from the given RCT is not equal to the ideal rate of the study population. Let $p_{RCT} = p(g(x) = 1|y' = 1)$ and $p_{IDEAL} = p(g(x) = 1|y = 1) = p(g(x) = 1)$.

We wish to test

$$H_0: p_{RCT} = p_{IDEAL}$$

$$H_A: p_{RCT} \neq p_{IDEAL}$$

$p_{RCT}$ is the observed rate of the subgroup in the given RCT, directly calculated from the actual RCT subject data.

$p_{IDEAL}$ is the ideal rate of the subgroup in the considered subpopulation. In this paper, $p_{IDEAL}$ is estimated using NHANES 2015-2016, a national survey that assesses the health and nutritional status of adults and children in the United States. Since NHANES uses a complex survey design, we estimate the ideal rate for each subgroup from NHANES data via the R package called "survey" using the provided survey weights in NHANES. For different categories of attributes, the selected survey weights are different. For example, for the data includes demographic attributes only, we use "wtint2yr" since all the information were obtained in the interview; for the data include lab results, subsample weights such as "wtmec2yr" were used to best represent the target population. Thus, the estimated ideal rates for a given attribute may vary slightly depending on the set of attributes selected.

Since we compare an observed proportion to an expected proportion for each subgroup, and there are only two categories (i.e. belong to the subgroup or not), we use the one-proportion z-test. This test is appropriate if the subgroup size is greater or equal to 5 and the number of subjects not in this subgroup is greater or equal to 5. Then, the test statistic has the formula $z = \frac{p_{RCT} - p_{IDEAL}}{\sigma} = \frac{p_{observed} - p_{IDEAL}}{\sqrt{\frac{p_{IDEAL}(1-p_{IDEAL})}{n_{RCT}}}}$, where $n_{RCT}$ represents the total number of participants in the RCT, and the standard deviation ($\sigma$) of the sampling distribution is $\sigma = \sqrt{\frac{p_{IDEAL}(1-p_{IDEAL})}{n_{RCT}}}$. Since we make multiple comparisons among subgroups, the Benjamini-Hochberg procedure is used to generate adjusted p-values (BH p-values) to control the false discovery rate. If the according adjusted p-value is smaller than the significance level $\alpha$, we cannot accept the null hypothesis, which means that the observed rate is significantly different from the ideal rate. For small subgroups, we use the exact binomial test to make the much more accurate since the normal approximation is only accurate for large sample sizes.

Some example statistical results are shown in Table S1 for Log Disparity for the ACCORD study. The full set of statistical results for all subgroups can be obtained from the Supplementary ZIP CSV file named Statistics Results.zip and they are also available on the GitHub repository, https://github.com/TheRensselaerIDEA/ClinicalTrialEquity.

**SUPPLEMENT 3: OTHER RESULTS**

All analysis results of available subgroups from ACCORD, ALLHAT, and SPRINT not shown in the paper can be obtained from our visualization tool implemented in R Shiny available at https://miao-qi-rpi-app.shinyapps.io/EquityBrowser. The interactive interface allows the user to construct representativeness visualization for any possible subgroups.

The R codes for the tool and analysis can be downloaded from the GitHub repository: https://github.com/TheRensselaerIDEA/ClinicalTrialEquity.

**SUPPLEMENT 4: RCT DATASETS**

The ACCORD study (1999 - 2009) randomly selected 10,251 adult subjects with type 2 diabetes mellitus from 77 clinical sites in North America. Its main purpose was to examine the effect of intensive treatment on three important CVD risk factors (i.e.

glycaemia, lipid, and blood pressure) in the prevention of major cardiovascular events in subjects with diabetes.

The ALLHAT study (1993-2002) randomly selected 33,357 hypertensive participants aged over 55 years from 623 North American centers in the hypertension study. Since we examined hypertension only in this case, the subjects for the lipid study of ALLHAT are excluded in our analysis. The objective of the hypertension study of ALLHAT is to compare the effectiveness of new treatments of blood pressure with diuretic-based treatments to reduce hypertension-related morbidity and mortality. ALLHAT featured a large sample size for hypertension study and multi-ethnic population.

The SPRINT study (2010-2016) randomly selected 9,361 participants aged over 50 with an SBP between 130 and 180 mm Hg and with at least one additional Cardiovascular disease (CVD) risk factor. Additionally, the enrollees should not have diabetes or a history of stroke. The SPRINT study tested whether the incidence of CVD can be reduced by an intensive antihypertensive treatment (target SBP <120 mm Hg) compared to a standard treatment (target SBP <140 mm Hg). Specifically, SPRINT recruitment had five target subgroups including minorities, women, subjects aged over 75, individuals with CVD, and individuals with chronic kidney disease.

In general, any subject attributes measured at baseline can be treated as protected attributes in our approach. However, considering the reliability of an estimate on the ideal frequency from NHANES, we eliminate self-reported attributes such as the rate of currently used antihypertensive medication. Additionally, the correlated attributes, such as systolic and diastolic blood pressures, are reduced to a single attribute (SBP).

All three RCTs are sponsored by the National Heart, Lung, and Blood Institute (NHLBI) and the data sets are available from https://biolincc.nhlbi.nih.gov.
